# Estimation of local time-varying reproduction numbers in noisy surveillance data

**DOI:** 10.1101/2021.04.23.21255958

**Authors:** Wenrui Li, Katia Bulekova, Brian Gregor, Laura F. White, Eric D. Kolaczyk

## Abstract

A valuable metric in understanding local infectious disease dynamics is the local time-varying reproduction number, i.e. the expected number of secondary local cases caused by each infected individual. Accurate estimation of this quantity requires distinguishing cases arising from local transmission from those imported from elsewhere. Realistically, we can expect identification of cases as local or imported to be imperfect. We study the propagation of such errors in estimation of the local time-varying reproduction number. In addition, we propose a Bayesian framework for estimation of the true local time-varying reproduction number when identification errors exist. And we illustrate the practical performance of our estimator through simulation studies and with outbreaks of COVID-19 in Hong Kong and Victoria, Australia.

## 2 Introduction

Epidemic modeling, while not at all new, has taken on renewed importance due to the COVID-19 pandemic. The local time-varying reproduction number is an important quantity to monitor the infectiousness and transmissibility of diseases and, therefore, to design and adjust public health responses during an outbreak. Recent examples include monitoring transmission of the COVID-19 pandemic and demonstrating the efficacy of non-pharmaceutical interventions in more than 100 countries [1–4]. The value of the local time-varying reproduction number, 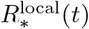, represents the expected number of secondary local cases arising from a primary case infected at time *t*. Different formal definitions of 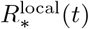 have been proposed, and a number of methods are available to estimate this quantity. The widely used EpiEstim estimator is an estimator of the instantaneous reproductive number that is defined as the ratio of the expected number of incident locally infected cases at time *t* to the expected total infectiousness of infected individuals at time *t* [5, 6]. In implementing this estimator, we typically smooth cases over a sliding window. This can have the result of making the estimator less timely but with the benefit of smoothing out much of the noise due to day of week effects in reporting and other random fluctuations to get a clearer trend.

Distinguishing local cases from imported cases is essential to estimation of the local time-varying reproduction number [5]. However, surveillance data generally is available only up to some level of error. For example, if we are unable to identify the correct source of infection from contact tracing or genetic information, imported cases might be misclassified as local cases, and vice versa. Such misclassification error is recognized as one limitation of estimating 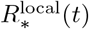 in the COVID-19 outbreak [7, 8]. We investigate how identification error impacts on the estimation of the instantaneous reproduction number and, thus, on our understanding of diseases transmission dynamics.

Extensive work regarding improving inference of time-varying reproduction numbers has been done. For instance, there have been efforts to estimate the serial interval that is used to compute the total infectiousness for 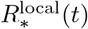 estimation, including Bayesian parametric estimation using data augmentation Markov Chain Monte Carlo [5, 9], and a cure model for limited follow-up data [10]. Many studies have explored the effects of imperfect detection and estimated the true infection prevalence [8, 11–13]. But, to our best knowledge, there has been little attention to date given towards accounting for identification errors of local and imported cases.

Our contribution in this paper is to quantify how such errors propagate to the local time-varying reproduction number, and to provide estimators for 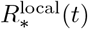 when contact tracing survey information is available. Adopting the definition of 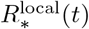 proposed by [5], we characterize the impact of identification errors on the bias of noisy local time-varying reproduction numbers. Our work shows that, in general, the bias can be expected to be nontrivial. Accordingly, we propose a Bayesian framework to estimate the true local time-varying reproduction number. Numerical simulation suggests that high accuracy is possible for estimating local time-varying reproduction numbers in outbreaks of even modest size. We illustrate the practical use of our estimators in the context of COVID-19 pandemic in Hong Kong and Victoria, Australia.

The organization of this paper is as follows. In Section 3 we show the bias of the noisy local time-varying reproduction number, and propose a Bayesian hierarchical framework to estimate the true local time-varying reproduction number with imperfect knowledge. Section 4 reports the practical performance of our estimators through simulation studies and with SARS-CoV-2 infections in Hong Kong and Australia. Finally, we conclude in Section 5 with a discussion of future directions for this work.

## 3 Methods

In this section, we first quantify the bias of the noisy local time-varying reproduction number when misidentification occurs in the surveillance data. We then build a Bayesian hierarchical framework to estimate true local time-varying reproduction numbers. We also propose a method to estimate misidentification rates based on contact tracing survey data, which informs the prior distribution in the model.

### 3.1 Notation

Both the seminal Fraser article [14] and the Thompson *et al*. article [5] we are working from use what seems a tendency in the epidemiology literature of conflating empirical processes and their means. From the perspective of designing the simulation study and including other Bayesian aspects, we necessarily distinguish between processes and means more precisely in this paper. Specifically, we use letter *I* to denote the empirical processes and letter *µ* to denote their means. The (local) time-varying reproduction number involves *µ* only. The plug-in estimator of the time-varying reproduction number in [14] involves *I* only. The estimator of the local time-varying reproduction number proposed by [5] involves both *µ* and *I*. One of the reasons that [5] used both empirical values and population values might be this estimator is easier to work with. We note that we use the sum notation for empirical processes and the integral notation for their means.

To clarify the terminology, we provide the technical differences among the terms error, bias and accuracy we used in the paper. If the surveillance data we have is not the same as the underlying truth, we say that the surveillance data is with some error. Here error implies the differences between the surveillance data and the truth. The bias of an estimator is the difference between this estimator’s expected value and the true value of the parameter being estimated. We say an estimator is of high accuracy if the bias and variance of the estimator are relatively small.

The number of newly infected cases at time *t, I*_*∗*_(*t*), is the sum of the numbers of local 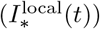 and imported 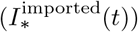 cases. If one assumes independence between calendar time and the generation interval, *g*(*s*), then the local time-varying reproduction number is defined as [5]

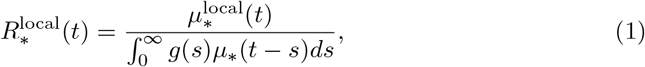

where 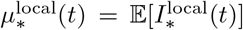 and *µ*_*∗*_(*t*) = 𝔼 [*I*_*∗*_(*t*)]. Note that from the perspective of simulation, the distinction between empirical values and population values seems potentially important, for the reason that “the expectation of a ratio is not the ratio of expectations”. Specifically, to calculate a true local time-varying reproduction number from simulation, we have expectations in the numerator and denominator, each of which can be approximated over a large number of trials through sample averages.

In reality, we only know the serial interval and the number of diagnosed cases. Let *I*(*t*), *I*^local^(*t*) and *I*^imported^(*t*) be the numbers of total diagnosed cases, local diagnosed cases, and imported diagnosed cases at time *t*, respectively. Then, we define a realistic local time-varying reproduction number as

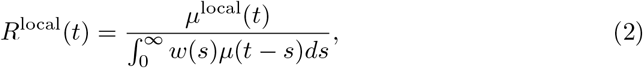

where *w*(*s*) is the serial interval, *µ*^local^(*t*) = 𝔼 [*I*^local^(*t*)] and *µ*(*t*) = 𝔼 [*I*(*t*)]. Note that the serial interval corresponds to date of symptom onset. One can estimate symptom onset dates by back calculation of report dates [15].

Realistically, we can expect identification of cases as local or imported to be imperfect. Let *Ĩ*^local^(*t*) and *Ĩ*^imported^(*t*) be the number of new local and imported cases reported at time *t*, with identification error. Thus, we define a noisy local time-varying reproduction number as

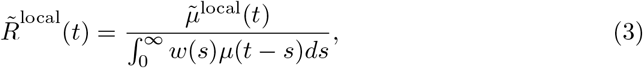

where 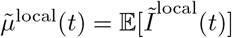.The definition of 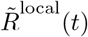 in (3) comes from an argument that mimics the original argument using Poisson arrivals in [14]. Specifically, we suppose that we observe a Poisson stream (also known as a Poisson process, i.e., a sequence of statistically independent and memoryless arrival times, the counts of which are Poisson distributed random variables) *Ĩ*^local^(*t*) that is a function of calendar time *t* in terms of the transmissibility, denoted 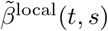, an arbitrary function of calendar time *t* and time since infection *s*. Then, 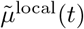 follows the so-called renewal equation

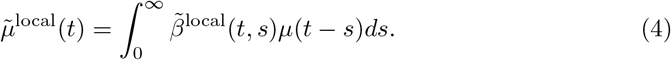

Following [14], we have

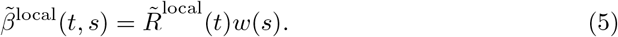

Inserting (5) into (4) yields the definition of 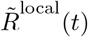 in (3).

Our interest is in characterizing the manner in which the uncertainty in *Ĩ*^local^(*t*) and *Ĩ*^imported^(*t*) propagates to the local time-varying reproduction number, and providing estimators of *R*^local^(*t*) to account for identification errors.

### 3.2 Bias of the noisy local time-varying reproduction number

We quantify the bias of the noisy local time-varying reproduction number in (3) when misidentification occurs. We begin by defining a model for *Ĩ*^local^(*t*) and *Ĩ*^imported^(*t*). Let *α*_0_ denote the probability that an imported case is misidentified as local, and *α*_1_ the probability that a local case is misidentified as imported. Then, a simple model is

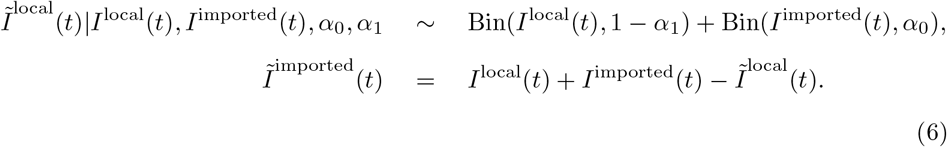

Under independence, the first relationship in (6) is directly obtained by the definition of *α*_0_ and *α*_1_. And the second equation in (6) is due to the fact that the total number of cases reported at time *t* is not affected by the misidentification.

By (6), the relationship between 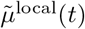 and *µ*^local^(*t*) is

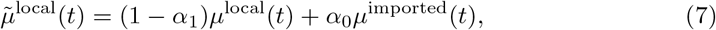

where *µ*^imported^(*t*) = 𝔼 (*I*^imported^(*t*)). Direct computation yields

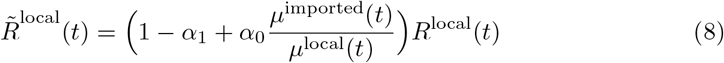

when *µ*^local^(*t*)≠0. From (8), we can see that the bias of 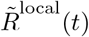 depends on *α, α* and the ratio of *µ*^imported^(*t*) and *µ*^local^(*t*). We will overestimate *R*^local^(*t*) if *α*_1_*/α*_0_ *< µ*^imported^(*t*)*/µ*^local^(*t*) and underestimate *R*^local^(*t*) if *α*_1_*/α*_0_ *> µ*^imported^(*t*)*/µ*^local^(*t*). The ratio of 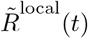 to *R*^local^(*t*) is shown below.

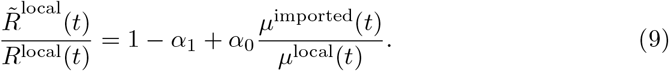

We can see that the ratio increases when *α*_0_ and *µ*^imported^(*t*)*/µ*^local^(*t*) increase, and decreases when *α*_1_ increases. The absolute difference of 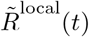 and *R*^local^(*t*) is as follows.

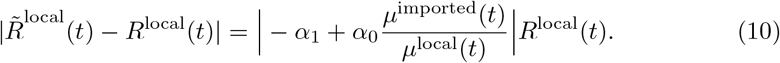

This absolute difference is proportional to *R*^local^(*t*) and the absolute difference of *α*_1_ and *α*_0_*µ*^imported^(*t*)*/µ*^local^(*t*).

### 3.3 Bayesian hierarchical modeling to account for misidentification

We propose a Bayesian framework to estimate *R*^local^(*t*) using noisy surveillance data. Figure 1 summarises the general idea.

**Fig 1.**
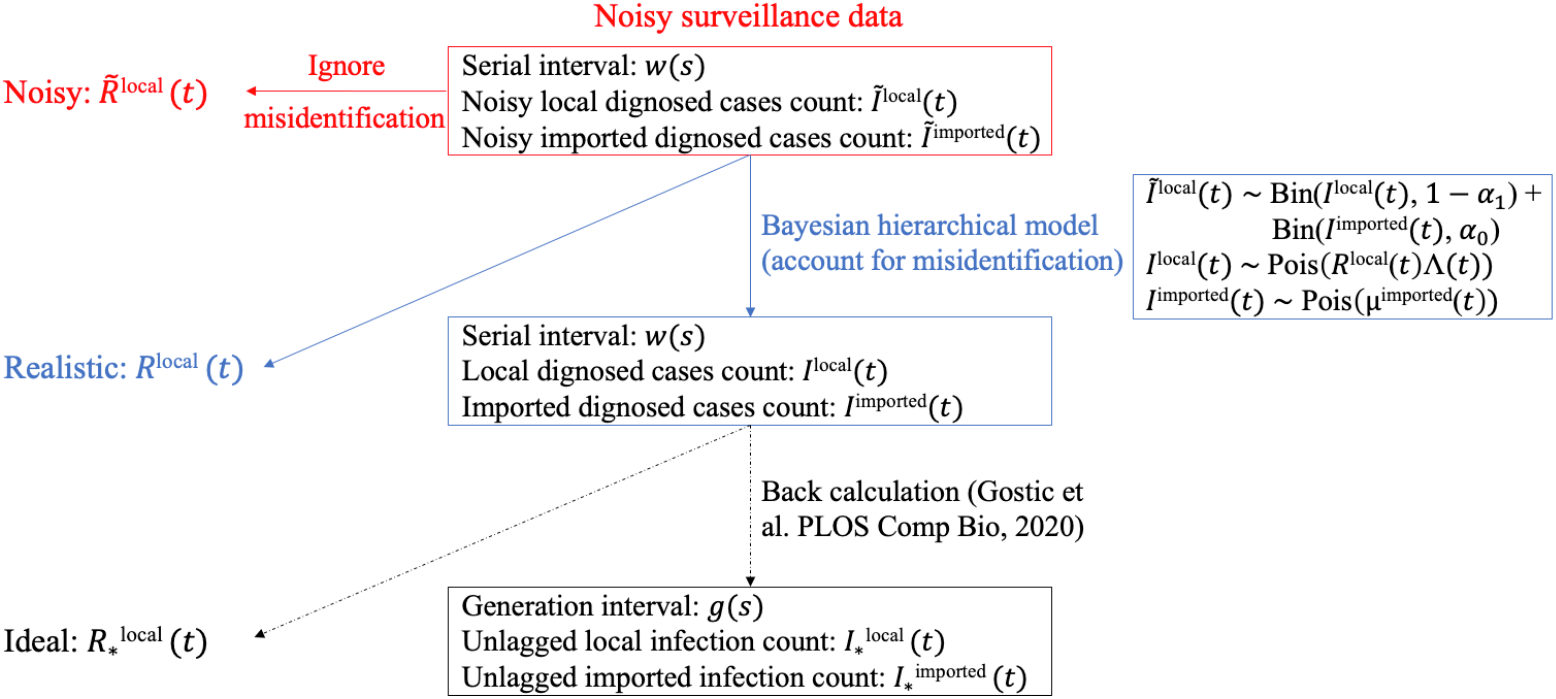
Schematic of our method to account for misidentification. Note that we do not back-calculate 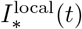 and 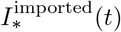 from estimated *I*^local^(*t*) and *I*^imported^(*t*) in this paper.

The model for the data *Ĩ*^local^(*t*) and *Ĩ*^imported^(*t*) is defined in (6). Following [5, 6, 14], we specify

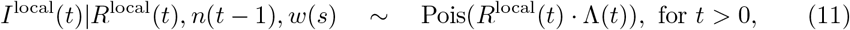

where 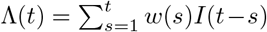 is the total infectiousness of infected individuals at time *t*, and *n*(*t*−1) represent the historical data up to time *t*−1 (i.e., *I*^local^(0), *I*^imported^(0), …, *I*^local^(*t*− 1), *I*^imported^(*t* − 1)). Note that Λ(*t*) is undefined for *t* = 0. So, we assume that

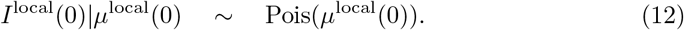

And we assume the imported case counts follow a Poisson distribution:

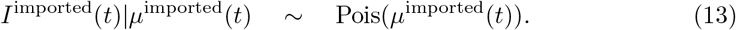

Next, we define relevant prior distributions. We assume a distribution for *R*^local^(*t*) of the form

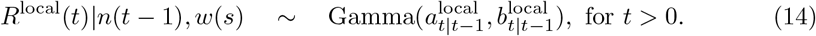

This choice is similar to that in [5], but differs in that we specify gamma conditioned on the history, rather than marginally. The conditioning reflects the expectation that the evolution of *R*^local^(*t*) is likely to depend on the course of infection in the population and intervention measures that may result. One can set 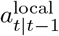 and 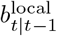 based on the historical surveillance data, e.g., 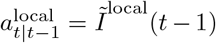 and 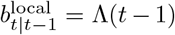. Analogously, we also assume gamma distributed priors for *µ*^imported^(*t*) and *µ*^local^(0), that is,

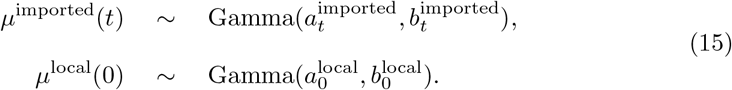

In addition, we assign the beta distributed priors to the misidentification rates:

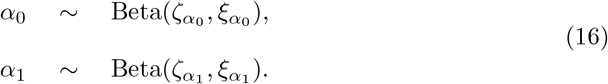

By using Markov chain Monte Carlo (MCMC) simulation, we can get both estimates of *R*^local^(*t*) and its uncertainty. We implement MCMC using the R package, NIMBLE [16–18] with the default assignment of sampler algorithms. The samplers assigned to the variables are as follows: Gibbs samplers are assigned to *µ*^local^(0) and *µ*^imported^(*t*), *t* ≥ 0, which have conjugate relationships between their prior distribution and the distributions of their stochastic dependents; slice samplers [19] are used for *I*^local^(*t*) and *I*^imported^(*t*), *t* ≥ 0; Metropolis-Hastings adaptive random-walk samplers are set to *α*_0_, *α*_1_ and *R*^local^(*t*), *t >* 0.

### 3.4 Setting hyperparameters and initial values in MCMC

Without any information on the misidentification rates, it is difficult to get an accurate estimator of *R*^local^(*t*). However, contact tracing data could provide adequate information to estimate the misidentification rates. Here we use contact tracing data to set informative priors on *α*_0_ and *α*_1_, and initial values of *I*^local^(*t*) and *I*^imported^(*t*).

Let *p*_*i*_ be the probability that we think individual *i* is a local case based on the survey. Then, *p*_*i*_ can be modeled as a mixture of *α*_0_ and 1 − *α*_1_. Note that 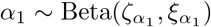 implies 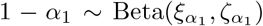. See the appendix for the proof of this property of the beta distribution. We thus model the distribution of *p*_*i*_ as a mixture of two beta distributions:

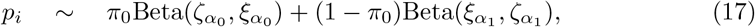

where *π*_0_ can be interpreted as the fraction of the diagnosed cases that are imported. By using an expectation–maximization (EM) algorithm, we can obtain estimators 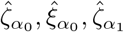 and 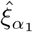. We set 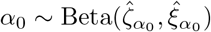 and 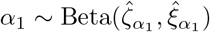 in the MCMC simulation.

Note that, if 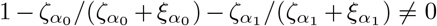, we obtain unbiased estimators of *I*^local^(*t*) and *I*^imported^(*t*)

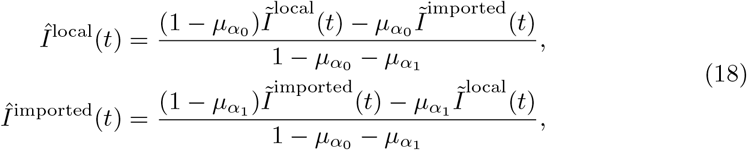

where 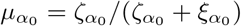 and 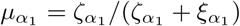. Thus, we set initial values of *I*^local^(*t*) and *I*^imported^(*t*) in the MCMC based on (18) and estimators 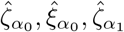 and 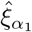. To be specific, the initial values of *I*^local^(*t*) and *I*^imported^(*t*) are given by

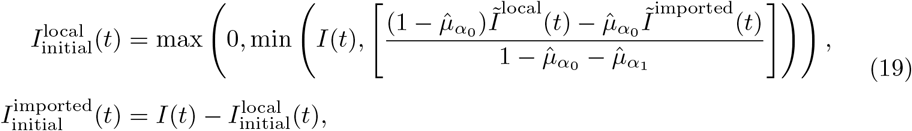

where 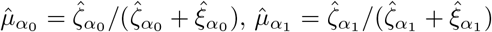, and [·] denotes the nearest integer. And we choose priors 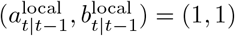 for *R*^local^(*t*), *µ*^imported^(*t*) ∼ Gamma(1, 1) and *µ*^local^(0) ∼ Gamma(1, 1), which are fairly uninformative.

## 4 Results

In this section, we conducted some simulations to illustrate the performance of the proposed estimation methods. And we applied our method to two real data sets. One is surveillance data of COVID-19 cases in Hong Kong that includes contact tracing information, including travel history data [20]. They collected information on 1,038 SARS-CoV-2 cases confirmed between 23 January and 28 April 2020. And they identified 355 local cases and 683 imported cases. The other data set is from the COVID-19 pandemic in Victoria, Australia, studied in [21]. There they had 1,333 laboratory-confirmed cases of COVID-19 between 6 January and 14 April 2020. After excluding duplicate patients from cases, they identified 345 local cases and 558 imported cases.

We considered two settings, a simulation setting and an application setting. In the simulation setting, we first used surveillance data from Hong Kong and Victoria to create realistic simulated data. Then, we added identification errors to the ‘true’ local and imported cases derived from the simulated epidemics. Finally, we estimated the local time-varying reproduction number using the noisy local and imported cases counts. In the application setting, we assumed that identified local and imported cases in the real data sets were with some error. The former results allow us to understand what properties can be expected of our estimators, while the latter are reflective of what would be observed in practice with such data.

### 4.1 Simulation study

In this simulation study, we used Covasim [22], a stochastic individual-based model for transmission of SARS-CoV-2, calibrated to the epidemics in Hong Kong and Victoria. In Covasim, a susceptible-exposed-infectious-removed (SEIR) model dictates the progression of disease for individuals, and contact networks determine interactions between individuals that can cause infection. Covasim supports an extensive set of interventions, including both non-pharmaceutical interventions and pharmaceutical interventions. In the calibration, we set network connectivity and intervention strategies such that the simulated data are close to the epidemics in Hong Kong and Victoria. The details of parameter values we used are available at https://github.com/KolaczykResearch/EstimLocalRt.

Figure 2 shows the average daily local and imported diagnosed counts over 1,000 trials. The noisy *Ĩ*^local^(*t*) and *Ĩ*^imported^(*t*) were generated according to (6). We set *α*_0_ ∼ 0.1 (beta distributed with mean 0.1), and *α*_1_ ∼ 0.3, 0.4 or 0.5 to see the effect of small *α*_0_ and large *α*_1_. This might happen if the definition of imported cases relies on travel history collected in the case investigation and some people are infected locally, even though they have a travel history within 14 days prior to symptom onset. We also considered *α*_1_ ∼ 0.1, and *α*_0_ ∼ 0.3, 0.4, or 0.5 (corresponding to small *α*_1_ and large *α*_0_, which might occur if cases are defined as local when we are not sure about their source of infection.) We assumed that both *α*_0_ and *α*_1_ are unknown.

**Fig 2.**
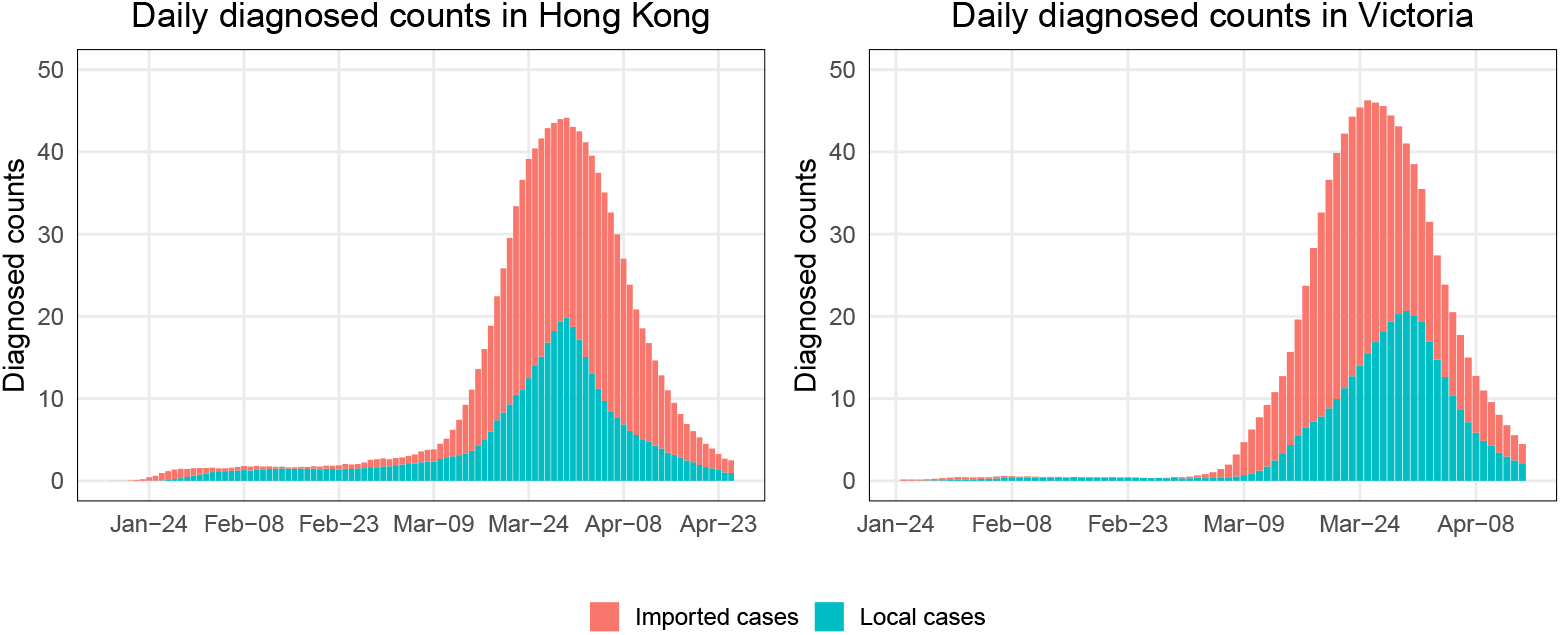
The means of daily local and imported diagnosed counts in 1,000 simulation trials for epidemics in Hong Kong and Victoria.

We evaluated the estimate for *R*^local^(*t*) in terms of a corresponding posterior, and 95% credible intervals. Figures 3 and 4 show the simulation results, in which we ran MCMC chains of 10,000 samples for each of 1,000 simulated epidemic trials. The number of burn-in samples is 1,000. And we used the trace and autocorrelation plots to evaluate the samples. In each trial, we compute the posterior mean and 95% credible intervals of estimated local time-varying reproduction numbers at each time point. Then we take the average over 1,000 trials and obtain the curves and error bands in Figures 3 and 4. Figure 3 assumes that we are more likely to misclassify local cases as imported cases and Figure 4 assumes that we are more likely to misclassify imported cases as local cases. The reason for not showing estimates for *R*^local^(*t*) in the left part of the right panel is that there are few diagnosed counts and the data are not sufficiently informative. The red curve represents the results obtained from our Bayesian model. For comparison purposes, we computed 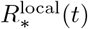 (corresponds to the blue curve) and *R*^local^(*t*) (corresponds to the purple curve) defined in (1) and (2) by approximating 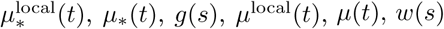 using 1,000 simulation trials. And we calculated the widely used estimator of 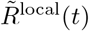 (corresponds to the green curve) defined in (3), which is implemented in the R package, EpiEstim [23]. We chose the weekly sliding window (default setting in EpiEstim) so the green curve has a thinner credible interval compared to the red curve. We view it as a representative estimator that does not account for misidentification, i.e., it treats the noisy local and imported cases as true. Note that the blue curve 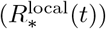 is temporally accurate. However, we used the lagged case observations and the serial interval in our Bayesian framework and EpiEstim. Thus, *R*^local^(*t*) (corresponds to the purple curve) is what we could estimate accurately using our Bayesian model.

**Fig 3.**
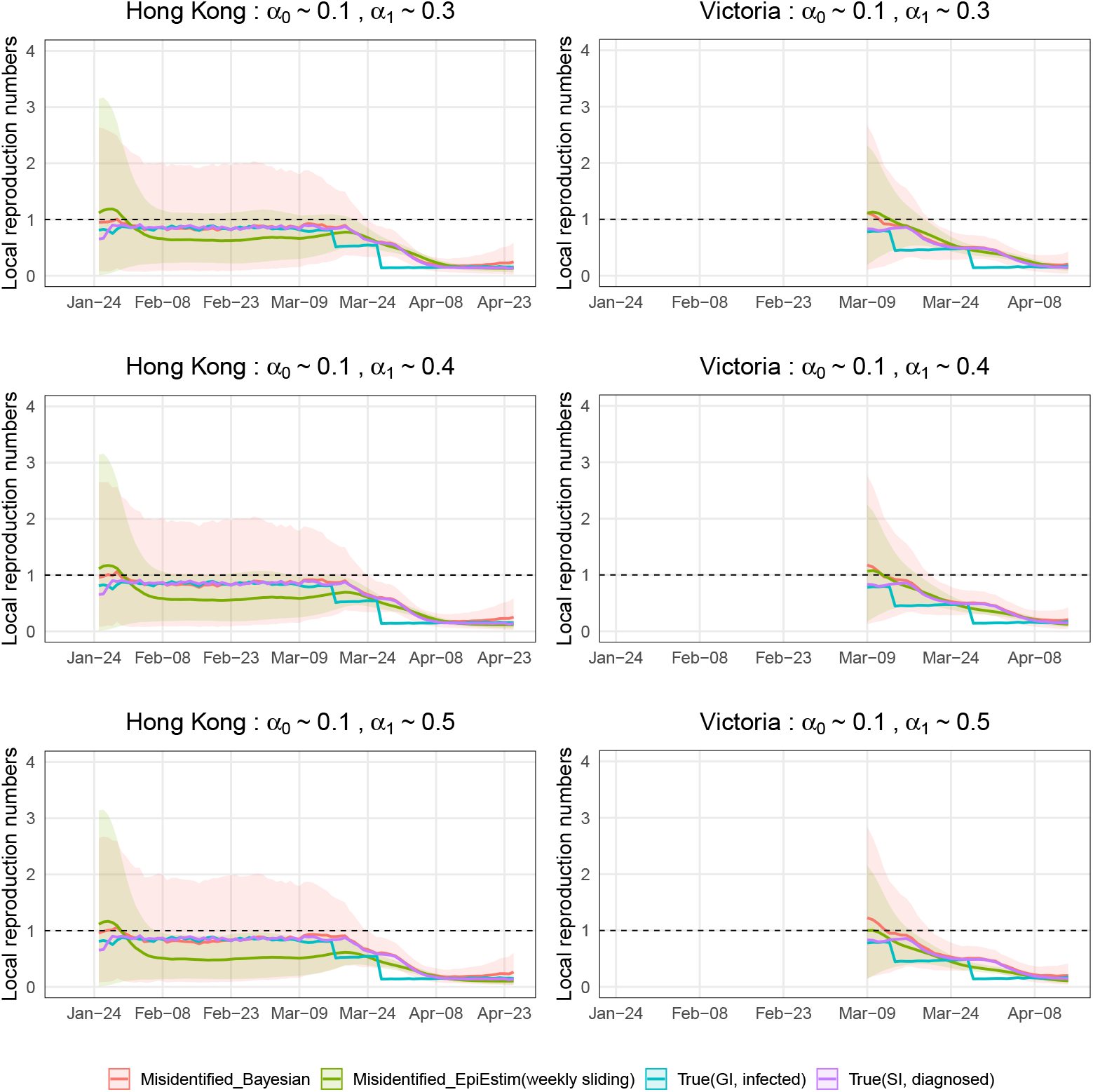
Estimations of local time-varying reproduction numbers in simulated epidemics for Hong Kong and Victoria under three sets of error misidentification rates: *α*_0_ ∼ 0.1, and *α*_1_∼ 0.3, 0.4, or 0.5. The error bands are the averages of 95% credible intervals over 1,000 trials at each time point.

**Fig 4.**
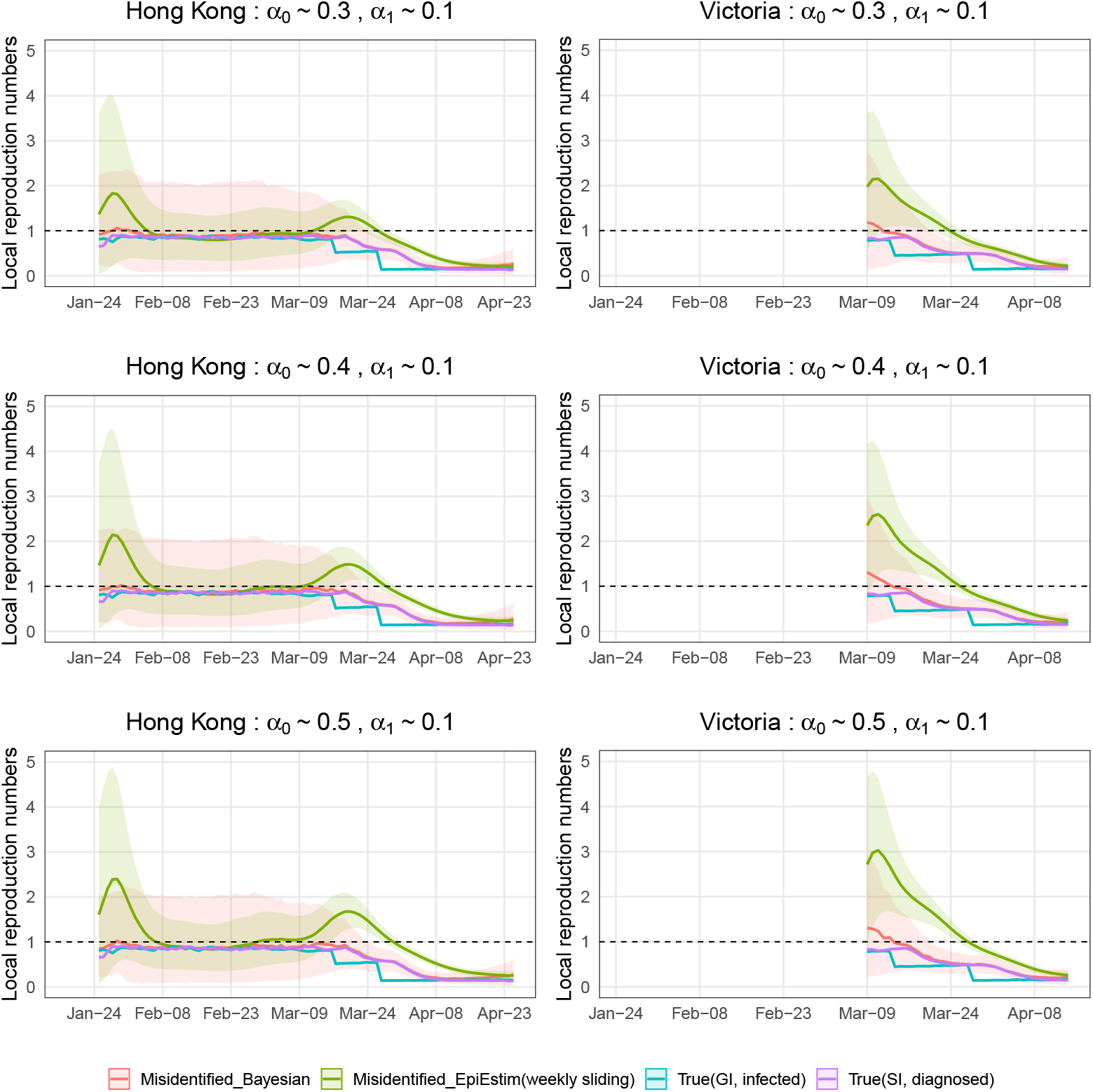
Estimations of local time-varying reproduction numbers in simulated epidemics for Hong Kong and Victoria under three sets of error misidentification rates: *α*_1_ ∼ 0.1, and *α*_0_ ∼ 0.3, 0.4, or 0.5. The error bands are the averages of 95% credible intervals over 1,000 trials at each time point.

Recall that the mean of unlagged infection counts was used in the blue curve 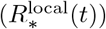 and the mean of lagged diagnosed cases counts was used in the purple curve (*R*^local^(*t*)). When the intervention strategy like shutting down is adopted (e.g., the middle of March in the simulated epidemic in Hong Kong), the infection counts will decrease sharply at the same time, but the diagnosed case counts will decrease smoothly with some time lag if we don’t test all people everyday. This is why we see sharp decreases in the blue curve and smooth decreases in the purple curve.

In the simulated epidemics for both Hong Kong and Victoria, if we ignore the misidentification, we will underestimate *R*^local^(*t*) when the mean of *α*_0_ is small and the mean of *α*_1_ is relatively large (Figure 3), and overestimate *R*^local^(*t*) when the mean of *α*_1_ is small and the mean of *α*_0_ is relatively large (Figure 4), with the biases increasing when the means of *α*_0_ and *α*_1_. The results are consistent with (8) implying that the biases will lead to inappropriate public health response, i.e., inadequate interventions or overreaction. We corrected the bias using our Bayesian hierarchical framework. The biases of our estimators are close to zero in all cases. The 95% credible intervals of our estimators are wide in the first two months because the number of incident cases are very low. For the last month or so when the diagnosed counts are relatively high, the 95% credible intervals are narrow.

### 4.2 Application

We applied our proposed methods to surveillance data of COVID-19 cases in Hong Kong and Victoria. Figures 5 (a) and (b) show the daily local and imported cases counts in Hong Kong and Victoria. For Hong Kong data, [20] calculated the serial intervals using a gamma distribution and estimated shape and rate parameters of 2.23 and 0.37, respectively (corresponding to a mean of around 6 days and standard deviation of around 4 days). There is no specific serial interval that has been calculated for Victoria. Considering the epidemic curve in Victoria is relatively similar to that in Hong Kong, we used the same serial interval distribution when we estimate *R*^local^(*t*) in Victoria.

**Fig 5.**
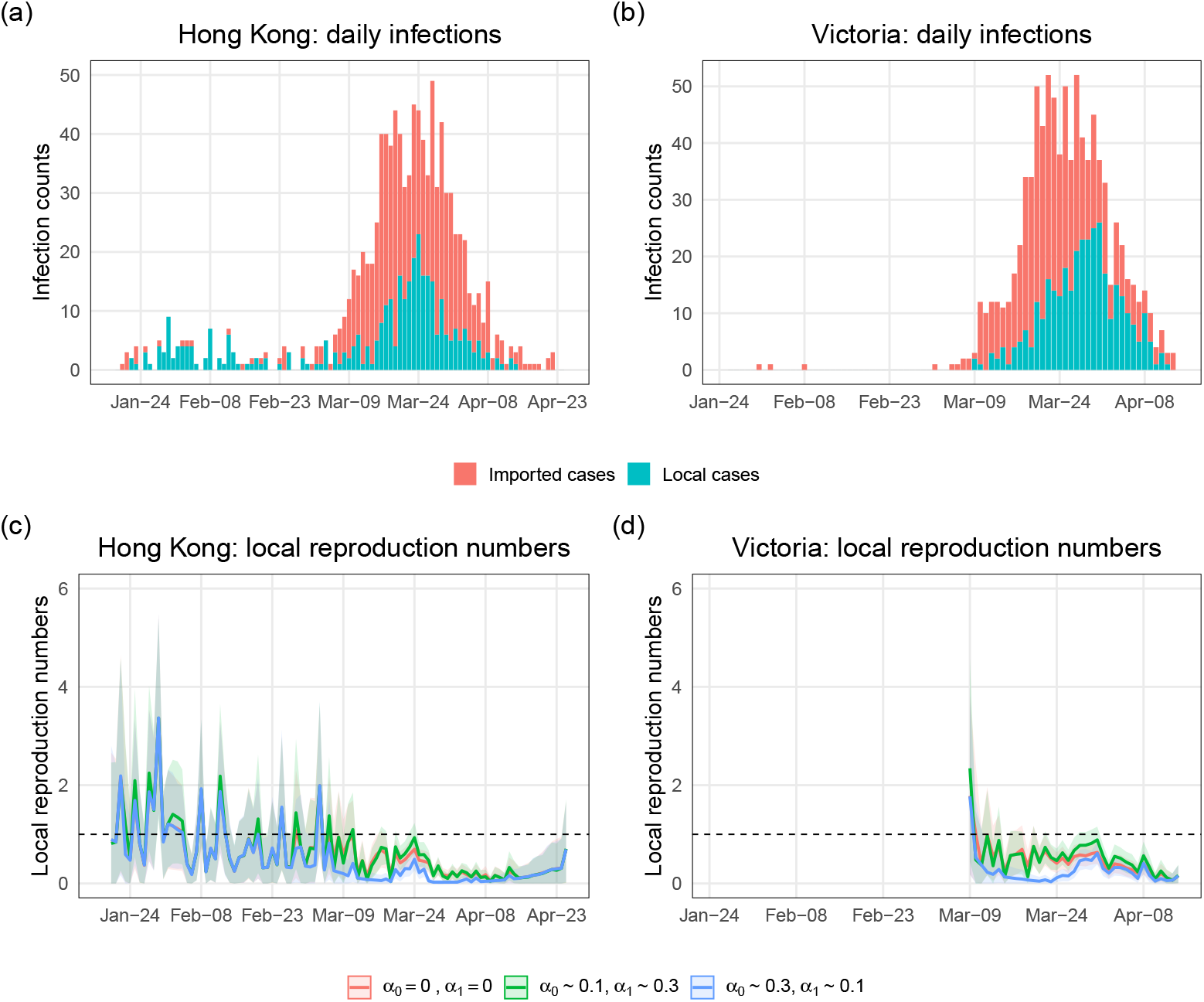
Epidemic curves of COVID-19 cases and estimations of local time-varying reproduction numbers in Hong Kong and Victoria. (a) The epidemic curve of daily cases of laboratory-confirmed SARS-CoV-2 infection in Hong Kong by symptom onset date and colored by case category. Asymptomatic cases are included here by date of confirmation. (b) The epidemic curve of the coronavirus disease cases in Victoria by sample collection date and colored by case category. (c) and (d) Estimations of local time-varying reproduction numbers under three assumed scenarios: 1) no identification error, 2) *α*_0_ ∼ 0.1 and *α*_1_ ∼ 0.3 (around 10% imported cases are misclassified as local and around 30% local cases are misclassified as imported), 3) *α*_0_ ∼ 0.3 and *α*_1_ ∼ 0.1 (around 30% imported cases are misclassified as local and around 10% local cases are misclassified as imported). The bands are the 95% credible intervals.

Since we did not have access to the contact tracing survey data mentioned in Section 2 3.4 to infer the misidentification rates, we investigated a range of plausible values. Figures 5 (c) and (d) show estimates for *R*^local^(*t*) under three assumed scenarios: 1) no identification error, 2) small *α*_0_ and large *α*_1_, 3) small *α*_1_ and large *α*_0_. We ran MCMC chains of 10,000 samples and the error bands are the 95% credible intervals. We can see that the estimated local time-varying reproduction numbers are quite different when the two identification error rates are about 10% and 30%. If we think we are more likely to misclassify local cases as imported, then we should trust the curve corresponding to scenario 2). If imported cases are more likely to be misidentified as local, then the curve corresponding to scenario 3) is reliable. And if we believe the identification error is close to zero, we should trust the estimate under scenario 1). For example, in late March, the estimated local time-varying reproduction numbers and 95% credible intervals are below one under scenario 1), but are near or above one under scenario 2). The differences can lead to different public health policies.

Ultimately, we see that the ability to account for identification error appropriately in reporting the local time-varying reproduction number can lead to substantially different conclusions than use of the original, noisy local time-varying reproduction number. These differences can then in turn be translated to decision making for public health response.

## 5 Discussion

We have developed a general framework for estimation of the true local time-varying reproduction numbers in contexts wherein one has identified local and imported case counts with some error. Simulations demonstrate that substantial inferential accuracy by our estimators is possible when nontrivial error is present. And our application to epidemics in Hong Kong and Victoria shows that the gains offered by our approach over presenting the noisy local instantaneous reproduction number can be pronounced.

We have shown examples on a state/province level, but our method could be useful for cities, or more local settings, such as a university trying to determine if there is substantial local transmission occurring. Our approach requires daily numbers of local and imported cases, serial interval, and contact tracing data or other data to provide adequate information to estimate the misidentification rates.

We have pursued a Bayesian approach to the problem of estimating the local instantaneous reproduction number. The credible intervals are relatively wide when the number of cases is low. To improve the performance at low case incidence, Kalman filtering is a natural approach. Estimating the time-vary reproduction number by Kalman filtering is an emerging topic. For instance, [24] constructed a recursive Bayesian smoother for estimating the effective reproduction number from the incidence of an infectious disease in real time and retrospectively. However, one typically does not distinguish between local and imported cases in this setting.

The identification errors are informed by contact tracing survey data in our approach. If the data from the survey is categorical (e.g., we ask people where they were infected and attach some qualitative measure of our confidence that we think they are local cases), we can transform them into numerical values. For example, [25] proposed a method that converts categorical variables to numerical data for a Gaussian distribution. We could modify the method to convert categorical variables to Beta distributed data. If the survey data is unavailable, using genomic data is a natural alternative. Genomic surveillance has been used to detect transmission clusters and to provide information on the possible source of individual cases [26–31].

We assume the identification errors are constant over time in our model. One future direction is relaxing this assumption. The identification errors may vary over time as the quality of surveillance data may not be the same. And the errors may depend on the incidence of local and imported cases. If there are few imported cases, an imported case might be likely to be incorrectly classified as local but a local case will be less likely to be incorrectly classified as imported.

We have shown the results of retrospective estimation. And it is computationally feasible to run MCMC on each day to obtain real time estimators; it takes about 5 minutes for the MCMC chain of 10,000 samples.

In the simulation study, we reported the mean of posterior means of estimated local time-varying reproduction numbers over 1,000 trials. To see if there is much variation in estimated values between simulations, we have computed the standard derivation of posterior means from 1,000 simulated epidemic trials at each time point. For the simulated epidemic in Hong Kong, the average of standard derivations (over time) is ranging from 0.37 to 0.43 in the six misidentification error scenarios shown in Figures 3 and 4. For the simulated epidemic in Victoria, the average of standard derivations (over time) is ranging from 0.28 to 0.38 in the six misidentification error scenarios shown in Figures 3 and 4.

We assume the serial interval for Victoria is the same as that in Hong Kong. There is variability in the serial interval among countries. [32] summarised 129 estimates of serial intervals reported for COVID-19, with means or medians ranging from 1.0 to 9.9. Also, serial interval observations for COVID-19 could be negative [33]. Exploring the robustness of our model to the serial interval could be a potential future direction.

The use of the lagged case observations and the serial interval can lead to temporal inaccuracies in the estimation of local time-varying reproduction numbers, which can hinder inference about the impact of changes in behavior and policies on the local transmission. The best practice is to back-calculate unlagged infection counts from lagged case observations [34]. Thus, to improve the accuracy of the estimation of local time-varying reproduction numbers, we can first back-calculate the unlagged infection counts using the noisy surveillance data and then run the MCMC with those unlagged counts.

If contact tracing datasets contain cases with unknown classification as local or imported, we could use the information from other data (e.g. genomic data) to impute these cases. If no other information is available, we could randomly classify these cases as local or imported.

As shown in the simulation study, ignoring misclassification of local or imported cases can lead to substantially inaccurate estimation of local time-varying reproduction numbers. In our data application, the misidentification rates are relatively small and thus the incorrect classification of local or imported cases does not have a big impact on the estimation of local time-varying reproduction numbers. However, there may be other real-world examples where our modeling framework becomes important.

While this paper was awaiting review, we became aware of related work that appeared by [35]. In that paper, those authors developed a Bayesian framework to estimate the local time-varying reproductive number, accounting for unlinked local cases and potential different infectiousness among local and imported cases. One of the main differences between their work and our work is that they assumed misspecification of the source of infection for local cases, but perfect classification of cases (i.e. *α*_0_ = *α*_1_ = 0).

## Data Availability

No primary data are used in this paper. Secondary data sources are taken from Adam, Dillon C., et al. “Clustering and superspreading potential of SARS-CoV-2 infections in Hong Kong.” Nature Medicine 26.11 (2020): 1714-1719. and Seemann, Torsten, et al. “Tracking the COVID-19 pandemic in Australia using genomics.” Nature communications 11.1 (2020): 1-9. These data and the code necessary to reproduce the results in this paper are available at https://github.com/KolaczykResearch/EstimLocalRt.

## Data Accessibility

No primary data are used in this paper. Secondary data sources are taken from [20, 21]. These data and the code necessary to reproduce the results in this paper are available at https://github.com/KolaczykResearch/EstimLocalRt.

